# Epidemiological and clinical features of 2019-nCoV acute respiratory disease cases in Chongqing municipality, China: a retrospective, descriptive, multiple-center study

**DOI:** 10.1101/2020.03.01.20029397

**Authors:** Di Qi, Xiaofeng Yan, Xumao Tang, Junnan Peng, Qian Yu, Longhua Feng, Guodan Yuan, An Zhang, Yaokai Chen, Jing Yuan, Xia Huang, Xianxiang Zhang, Peng Hu, Yuyan Song, Chunfang Qian, Qiangzhong Sun, Daoxin Wang, Jin Tong, Jianglin Xiang

## Abstract

**Background:** In January 19, 2020, first case of 2019 novel coronavirus (2019-nCoV) pneumonia (COVID-19) was confirmed in Chongqing municipality, China.

**Methods:** In this retrospective, descriptive, multiple-center study, total of 267 patients with COVID-19 confirmed by real-time RT-PCR in Chongqing from Jan 19 to Feb 16, 2020 were recruited. Epidemiological, demographic, clinical, radiological characteristics, laboratory examinations, and treatment regimens were collected on admission. Clinical outcomes were followed up until Feb 16, 2020.

**Results:** 267 laboratory-confirmed COVID-19 patients admitted to 3 designated-hospitals in Chongqing provincial municipality from January 19 to February 16, 2020 were enrolled and categorized on admission. 217 (81.27%) and 50 (18.73%) patients were categorized into non-severe and severe subgroups, respectively. The median age of patients was 48.0 years (IQR, 35.0-65.0), with 129 (48.3%) of the patients were more than 50 years of age. 149 (55.8%) patients were men. Severe patients were significantly older (median age, 71.5 years [IQR, 65.8-77.0] *vs* 43.0 years [IQR, 32.5-57.0]) and more likely to be male (110 [50.7%] *vs* 39 [78.0%]) and have coexisting disorders (15 [30.0%] *vs* 26 [12.0%]). 41 (15.4%) patients had a recent travel to Hubei province, and 139 (52.1%) patients had a history of contact with patients from Hubei. On admission, the most common symptoms of COVID-19 were fever 225(84.3%), fatigue (208 [77.9%]), dry cough (189 [70.8%]), myalgia or arthralgia (136 [50.9%]). Severe patients were more likely to present dyspnea (17 [34.0%] *vs* 26 [12.0%]) and confusion (10 [20.0%] *vs* 15 [6.9%]). Rales (32 [12.0%]) and wheezes (20 [7.5%]) are not common noted for COVID-19 patients, especially for the non-severe (11 [5.1%], 10 [4.6%]). 118 (44.2%). Most severe patients demonstrated more laboratory abnormalities. 231 (86.5%), 61 (22.8%) patients had lymphopenia, leukopenia and thrombocytopenia, respectively. CD4^+^T cell counts decrease was observed in 77.1 % of cases, especially in the severe patients (45, 100%). 53.1% patients had decreased CD^+^3 T cell counts, count of CD8^+^T cells was lower than the normal range in part of patients (34.4%). More severe patients had lower level of CD4^+^ T cells and CD^+^3 T cells (45 [100.0%] *vs* 29[56.9%], 31 [68.9%] *vs* 20 [39.2%]). Most patients had normal level of IL-2, IL-4, TNF-α and INF-γ, while high level of IL-6 and IL-17A was common in COVID-19 patients (47 [70.1%], 35 [52.2%]). Level of IL-6, IL-17A and TNF-α was remarkably elevated in severe patients (32 [84.2%] *vs* 15 [51.7%], 25 [65.8%] *vs* 10 [34.5%], 17 [44.7%] *vs* 5 [17.2%]). All patients received antiviral therapy (267, 100%). A portion of severe patients (38, 76.0%) received systemic corticosteroid therapy. Invasive mechanical ventilation in prone position, non-invasive mechanical ventilation, high-flow nasal cannula oxygen therapy was adopted only in severe patients with respiratory failure (5[10.0%], 35[70.0%], 12[24.0%]). Traditional Chinese medicine was adopted to most of severe patients (43,86.0%).

**Conclusion:** Our study firstly demonstrated the regional disparity of COVID-19 in Chongqing municipality and further thoroughly compared the differences between severe and non-severe patients. The 28-day mortality of COVID-19 patients from 3 designed hospitals of Chongqing is 1.5%, lower than that of Hubei province and mainland China including Hubei province. However, the 28-mortality of severe patients was relatively high, with much higher when complications occurred. Notably, the 28-mortality of critically severe patients complicated with severe ARDS is considerably as high as 44.4%. Therefore, early diagnosis and intensive care of critically severe COVID-19 cases, especially those combined with ARDS, will be considerably essential to reduce mortality.

## Introduction

Since December of 2019, a cluster of cases of pneumonia with unknown etiology occurred in Wuhan, Hubei Province, China^1^. A novel coronavirus named the 2019 novel coronavirus (2019-nCoV) was soon isolated on January 12, 2020 and identified as the causative pathogen of this pneumonia^2^. On February 21, 2020, National Health Commission of the republic of China re-named this 2019-nCo-infected pneumonia as 2019-nCoV acute respiratory disease (COVID-19). Person-to-person transmission of COVID-19 in hospital and family settings is reported to be accumulating^3^. As of February 16, 2020, there are 70548 laboratory-confirmed cases and 1770 death cases in China, including 551 confirmed cases in Chongqing. The first laboratory-confirmed case of COVID-19 in the Chongqing municipality is reported on January 19, 2020.

Coronaviruses (CoV) are enveloped non-segmented positive-sense RNA viruses belonging to the family Coronaviridae, which can be transmitted between animals and humans. The 2019-nCoV is the seventh member of enveloped RNA coronavirus^2^. Coronavirus infections cause illness ranging from the mild cold to severe respiratory diseases such as Middle East Respiratory Syndrome (MERS-CoV) and Severe Acute Respiratory Syndrome (SARS-CoV), with mortality rates of 10% for SARS-CoV and 37% for MERS-CoV^4^. So far, a few patients of COVID-19 have developed severe pneumonia, acute respiratory distress syndrome (ARDS), multiple organ dysfunction syndrome (MODS) and had died^5^.

Recently, there are some studies demonstrating the clinical characteristics and epidemiology of COVID-19 patients in Wuhan or throughout China^5-8^. As Chongqing is reported to be one of the major cities imported passengers from Wuhan in China from December 16, 2019^9^. Given the regional disparity of COVID-19, we aim to describe epidemiological, clinical, laboratory, and radiological features, treatment, and prognosis of COVID-19 patients in Chongqing municipality, and compare the differences between non-severe and severe patients, which may unravel risk factors associated with 28-day mortality and further suggest a specific therapeutic intervention for patients with COVID-19 in Chongqing municipality, China.

## Methods

### Study Design and patients

A retrospective, descriptive, multiple-center study on the clinical characteristics of laboratory-confirmed cases with COVID-19 in Chongqing was performed in accordance with the Declaration of Helsinki. 267 laboratory-confirmed COVID-19 patients admitted to 3 designated-hospitals (Qianjiang central hospital of Chongqing, Chongqing three gorges central hospital and Chongqing public health medical center) in Chongqing provincial municipality from January 19 to February 16, 2020 were enrolled and categorized on admission. Patients with COVID-19 were diagnosed based on the World Health Organization (WHO) interim guidance^10^ and categorized into severe and non-severe COVID-19 according to the American Thoracic Society guideline. ARDS was defined according to the Berlin definition^11^. Informed consent was yielded due to the anonymous analysis of clinical data in retrospective study. This research was approved by the institutional ethics board of the Second Affiliated Hospital of Chongqing Medical University (No.2020-09), Chongqing public health medical center (No.2020-015-01-KY), Chongqing three gorges central hospital (No.2020-13) and Qianjiang central hospital of Chongqing (No.2020-07).

### Sample collection

Respiratory specimens, including nasopharyngeal swab and bronchoalveolar lavage fluid (BALF), or anal swab specimens were collected at admission to detect the presence of 2019-nCoV by real-time reverse-transcriptase polymerase-chain-reaction (RT-PCR) assay in designated authoritative laboratories of local centers for disease control and prevention^12^. Bacterial, fungal, and pathogenic microorganism test in respiratory and blood samples was routinely performed. Cytokines and lymphocyte subsets in peripheral blood were measured by fluorescence-labeled flow cytometry (Beckman, Cell Lab Quanta SC)

### Data Collection

The epidemiological, demographic, clinical, laboratory, radiological, treatment and outcomes data from confirmed COVID-19 patients’ medical and nursing records were obtained and analyzed. The date of disease onset was defined as the day when the symptoms were noticed. Clinical outcomes were followed up to February 16, 2020. Data were entered into a computerized database and were checked by two physicians.

### Statistical analysis

Continuous variables were expressed as the means and standard deviations (SD) or medians and interquartile ranges (IQR) as appropriate. Categorical variables were presented as the counts and percentages. Independent group t tests were used for comparison of means for continuous variables when the data were normally distributed; otherwise, the Mann-Whitney U test was used. Proportions for categorical variables were compared using the χ2 test or Fisher exact test. Data analyses were performed using GraphPad Prism 7.0 software and SPSS 19.0 software. A value of *p*< 0.05 was considered statistically significant.

## Results

### Epidemiology and clinical characteristics

Of all 309 patients enrolled as of February 16, 2020, 42 patients were excluded due to incompleteness of crucial data in original reports. None of medical staff were infected. Hence this study includes 267 patients with COVID-19 from 3 designed-hospitals in Chongqing municipality, China.

The epidemiological and clinical characteristics are shown in Table 1. The median age of the patients was 48.0 years (IQR, 35.0-65.0), with 129 (48.3%) of the patients were more than 50 years of age. 149 (55.8%) patients were men. Most of cases (80.1%) had no smoke history. In our study, only 41(15.4%) patients had underlying comorbidities including overweight and obesity (30 [11.2%]), diabetes (26 [9.7%]), respiratory system disease diseases (25 [9.4%]), hypertension (20 [7.5%]), cardiovascular and cerebrovascular disease (13 [4.9%]), digestive system disease (12 [4.5%]) and malignant tumor (2 [0.7%]). 41 (15.4%) COVID-19 patients had a recent travel to Wuhan or Hubei province, 139 (52.1%) patients had a history of contact with patients from Wuhan or Hubei, 18 (6.7%) patients had definite contact with the patients infected by confirmed COVID-19 patients from Wuhan or Hubei province. No definite epidemiological link was found in the 69 (25.8%) patients. There is only one patient in our study had definite direct exposure to Huanan seafood market.

**Table 1.** Epidemiological and clinical characteristics of COVID-19 patients in Chongqing.

| Clinical characteristics, symptoms or signs | Disease severity |  |  |  |
| --- | --- | --- | --- | --- |
|  | All patients<br>(n=267) | Non-severe<br>(n=217) | Severe<br>(n=50) | P value |
| Age, Median (range) – yrs | 48.0(20-80) | 43.0(20-79) | 71.5(39-80) |  |
| Age, Median (IQR) – yrs | 48.0(35.0-65.0) | 43.0(32.5-57.0) | 71.5(65.8-77.0) | < 0.001 |
| Age groups – No. % |  |  |  |  |
| < 50 yrs | 138(51.7) | 136(62.7) | 2(4.0) | < 0.001 |
| ≥ 50 yrs | 129(48.3) | 81(37.3) | 48(96.0) | - |
| Male sex – No., % | 149(55.8) | 110(50.7) | 39(78.0) | < 0.001 |
| Smoking history – No., % |  |  |  |  |
| smokers | 53(19.9) | 22(10.1) | 31(62.0) | < 0.001 |
| Non-smokers | 214(80.1) | 195(89.9) | 19(38.0) | - |
| Comorbidities – No., % |  |  |  |  |
| Any | 41(15.4) | 26(12.0) | 15(30.0) | 0.001 |
| Overweight or obesity | 30(11.2) | 16(7.4) | 14(28.0) | < 0.001 |
| Diabetes | 26(9.7) | 14(6.5) | 12(24.0) | < 0.001 |
| Hypertension | 20(7.5) | 7(3.2) | 13(26.0) | < 0.001 |
| Respiratory system disease | 25(9.4) | 15(6.9) | 10(20.0) | 0.009 |
| Cardiovascular and cerebrovascular diseases | 13(4.9) | 9(4.1) | 4(8.0) | 0.437 |
| Exposure to source of transmission within 14 days – No., % |  |  |  |  |
| a recent travel to Wuhan or Hubei province | 41(15.4) | 30(13.8) | 11(22.0) | 0.148 |
| contact with patients from Wuhan or Hubei | 139(52.1) | 113(52.1) | 26(52.0) | 0.992 |
| contact with the patients infected by confirmed patients from Wuhan. | 18(6.7) | 16(7.4) | 2(4.0) | 0.540 |
| No definite epidemiological link | 69(25.8) | 58(26.7) | 11(22.0) | 0.592 |
| Symptoms and signs |  |  |  |  |
| Respiratory symptoms – No., % |  |  |  |  |
| Fever on admission | 225(84.3) | 186(85.7) | 39(78.0) | 0.197 |
| Temperature on admission (°C) | 37.6(36.1-40.4) | 37.6(36.1-40.4) | 37.5(36.5-40.2) | 0.205 |
| 37.3-38.0 | 181(67.8) | 151(69.6) | 30(60.0) | 0.191 |
| > 38.0 | 44(16.5) | 35(16.1) | 9(18.0) | 0.748 |
| myalgia or arthralgia | 136(50.9) | 99(45.6) | 37(74.0) | < 0.001 |
| Cough | 189(70.8) | 155(71.4) | 34(68.0) | 0.631 |
| Fatigue | 208(77.9) | 170(78.3) | 38(76.0) | 0.719 |
| Chill | 30(11.2) | 19(8.8) | 11(22.0) | 0.008 |
| Nasal congestion | 53(19.9) | 44(20.3) | 9(18.0) | 0.716 |
| pharyngalgia | 41(15.4) | 33(15.2) | 8(16.0) | 0.889 |
| Hemoptysis | 5(1.9) | 3(1.4) | 2(4.0) | 0.236 |
| Dyspnea | 43(16.1) | 26(12.0) | 17(34.0) | < 0.001 |
| confusion | 25(9.4) | 15(6.9) | 10(20.0) | 0.009 |
| Nausea or vomiting | 6(2.2) | 5(2.3) | 1(2.0) | 1.000 |
| Diarrhea | 10(3.7) | 7(3.2) | 3(6.0) | 0.604 |
| anorexia | 46(17.2) | 33(15.2) | 13(26.0) | 0.094 |
| **Signs – No., %** |  |  |  |  |
| Throat congestion | 6(2.2) | 4(1.8) | 2(4.0) | 0.690 |
| Tonsil swelling | 3(1.1) | 2(0.9) | 1(2.0) | 0.465 |
| Enlargement of lymph nodes | 3(1.1) | 2(0.9) | 1(2.0) | 0.465 |
| rales | 32(12.0) | 11(5.1) | 21 (42.0) | < 0.001 |
| wheezes | 20(7.5) | 10(4.6) | 10(20.0) | 0.001 |
| **Onset of symptom to, median (IQR), d** |  |  |  |  |
| diagnosis | 4(2.0-8.0) | 4(2.0-7.0) | 7(3.0-11.7<br>5) | < 0.001 |
| Hospital admission | 7(3.0-10.0) | 6(3.0-9.0) | 9(6.0-12.7<br>5) | < 0.001 |

On admission, the most common symptoms of COVID-19 were fever 225(84.3%), fatigue (208 [77.9%]), dry cough (189 [70.8%]), myalgia or arthralgia (136 [50.9%]). Less common initial symptoms included nasal congestion (53 [19.9%]), pharyngalgia (41 [15.4%]), dyspnea (43 [16.1%]), chill (30 [11.2%]), confusion (25 [9.4%]). A portion of patients initially presented with symptoms of digestive system such as anorexia (46 [17.2%]), diarrhea (10 [3.7%]) nausea or vomiting (6 [2.2%]).

Abnormal signs of physical examination on admission, including throat congestion (6 [2.2%]), tonsil swelling (3 [1.1%]) and enlargement of lymph nodes (3 [1.1%]), are relatively rare. Notably, for auscultation of abnormal breath sound, rales (32 [12.0%]) and wheezes (20 [7.5%]) are not common noted for COVID-19 patients in Chongqing, especially for the non-severe patients (11 [5.1%], 10 [4.6%]). The median durations from symptoms onset to laboratory diagnosis and hospital admission were 4 days (IQR, 2.0-8.0) and 7 days (IQR, 3.0-10.0) respectively.

### Laboratory and radiologic results

The Laboratory and radiologic findings are shown in Table 2. On day of hospital admission, most patients had normal level of leucocytes (134 [50.2%]) and neutrophils (146 [54.7%]) counts, with 118 (44.2%), 231 (86.5%), 61 (22.8%) patients had lymphopenia, leukopenia and thrombocytopenia, respectively.

**Table 2.**
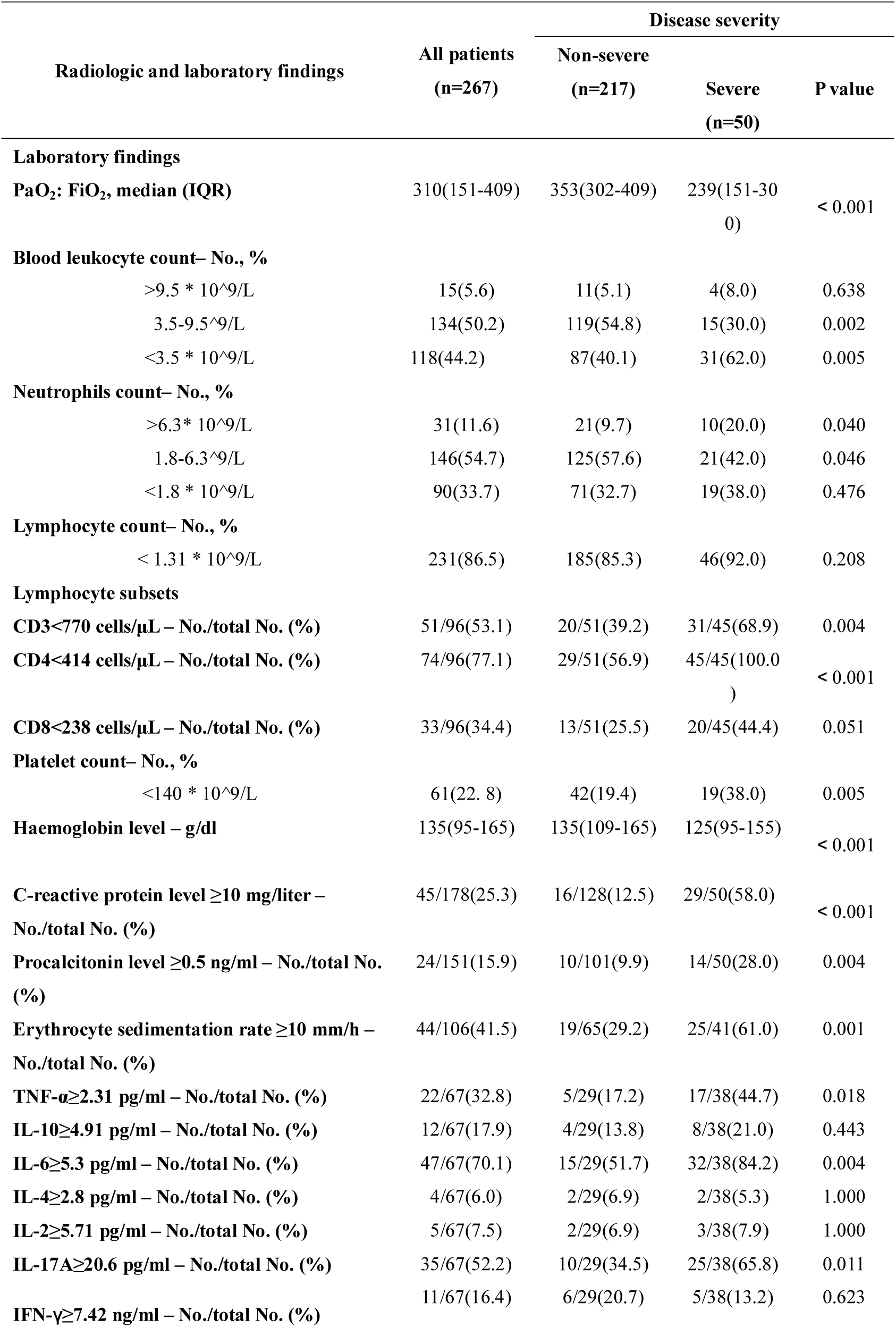
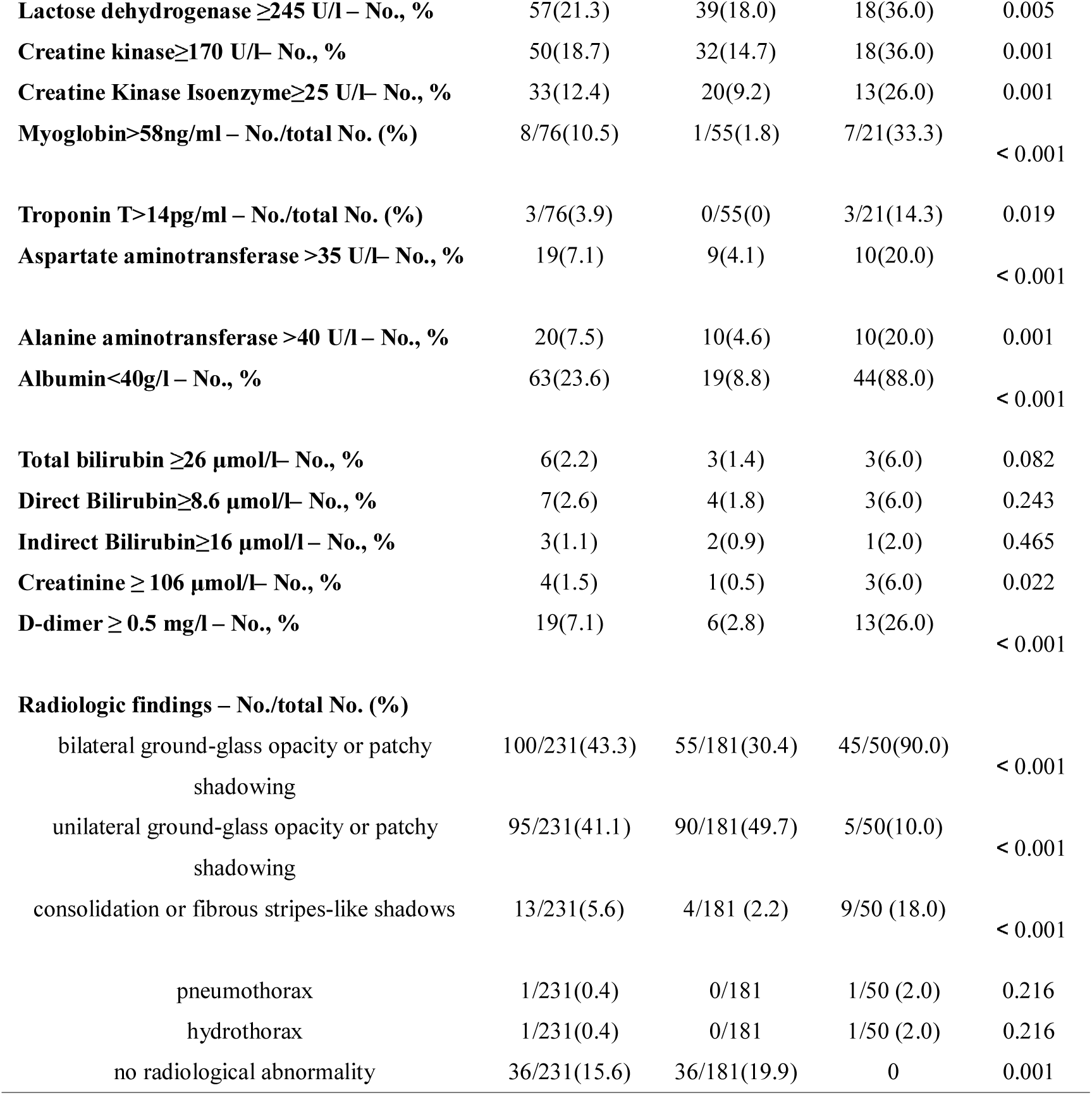
Laboratory and radiographic and findings of COVID-19 patients in Chongqing.

| Radiologic and laboratory findings | All patients<br>(n=267) | Disease severity |  |  |
| --- | --- | --- | --- | --- |
|  |  | Non-severe<br>(n=217) | Severe<br>(n=50) | P value |
| Laboratory findings |  |  |  |  |
| PaO<sub>2</sub>: FiO<sub>2</sub>, median (IQR) | 310(151-409) | 353(302-409) | 239(151-300) | < 0.001 |
| Blood leukocyte count– No., % |  |  |  |  |
| >9.5 * 10^9/L | 15(5.6) | 11(5.1) | 4(8.0) | 0.638 |
| 3.5-9.5^9/L | 134(50.2) | 119(54.8) | 15(30.0) | 0.002 |
| <3.5 * 10^9/L | 118(44.2) | 87(40.1) | 31(62.0) | 0.005 |
| Neutrophils count– No., % |  |  |  |  |
| >6.3* 10^9/L | 31(11.6) | 21(9.7) | 10(20.0) | 0.040 |
| 1.8-6.3^9/L | 146(54.7) | 125(57.6) | 21(42.0) | 0.046 |
| <1.8 * 10^9/L | 90(33.7) | 71(32.7) | 19(38.0) | 0.476 |
| Lymphocyte count– No., % |  |  |  |  |
| < 1.31 * 10^9/L | 231(86.5) | 185(85.3) | 46(92.0) | 0.208 |
| Lymphocyte subsets |  |  |  |  |
| CD3<770 cells/μL – No./total No. (%) | 51/96(53.1) | 20/51(39.2) | 31/45(68.9) | 0.004 |
| CD4<414 cells/μL – No./total No. (%) | 74/96(77.1) | 29/51(56.9) | 45/45(100.0) | < 0.001 |
| CD8<238 cells/μL – No./total No. (%) | 33/96(34.4) | 13/51(25.5) | 20/45(44.4) | 0.051 |
| Platelet count– No., % |  |  |  |  |
| <140 * 10^9/L | 61(22.8) | 42(19.4) | 19(38.0) | 0.005 |
| Haemoglobin level – g/dl | 135(95-165) | 135(109-165) | 125(95-155) | < 0.001 |
| C-reactive protein level ≥10 mg/liter – No./total No. (%) | 45/178(25.3) | 16/128(12.5) | 29/50(58.0) | < 0.001 |
| Procalcitonin level ≥0.5 ng/ml – No./total No. (%) | 24/151(15.9) | 10/101(9.9) | 14/50(28.0) | 0.004 |
| Erythrocyte sedimentation rate ≥10 mm/h – No./total No. (%) | 44/106(41.5) | 19/65(29.2) | 25/41(61.0) | 0.001 |
| TNF-α≥2.31 pg/ml – No./total No. (%) | 22/67(32.8) | 5/29(17.2) | 17/38(44.7) | 0.018 |
| IL-10≥4.91 pg/ml – No./total No. (%) | 12/67(17.9) | 4/29(13.8) | 8/38(21.0) | 0.443 |
| IL-6≥5.3 pg/ml – No./total No. (%) | 47/67(70.1) | 15/29(51.7) | 32/38(84.2) | 0.004 |
| IL-4≥2.8 pg/ml – No./total No. (%) | 4/67(6.0) | 2/29(6.9) | 2/38(5.3) | 1.000 |
| IL-2≥5.71 pg/ml – No./total No. (%) | 5/67(7.5) | 2/29(6.9) | 3/38(7.9) | 1.000 |
| IL-17A≥20.6 pg/ml – No./total No. (%) | 35/67(52.2) | 10/29(34.5) | 25/38(65.8) | 0.011 |
| IFN-γ≥7.42 ng/ml – No./total No. (%) | 11/67(16.4) | 6/29(20.7) | 5/38(13.2) | 0.623 |
| **Lactose dehydrogenase $\geq 245$ U/l – No., %** | 57(21.3) | 39(18.0) | 18(36.0) | 0.005 |
| **Creatine kinase $\geq 170$ U/l– No., %** | 50(18.7) | 32(14.7) | 18(36.0) | 0.001 |
| **Creatine Kinase Isoenzyme $\geq 25$ U/l– No., %** | 33(12.4) | 20(9.2) | 13(26.0) | 0.001 |
| **Myoglobin $> 58$ ng/ml – No./total No. (%)** | 8/76(10.5) | 1/55(1.8) | 7/21(33.3) | $< 0.001$ |
| **Troponin T $> 14$ pg/ml – No./total No. (%)** | 3/76(3.9) | 0/55(0) | 3/21(14.3) | 0.019 |
| **Aspartate aminotransferase $> 35$ U/l– No., %** | 19(7.1) | 9(4.1) | 10(20.0) | $< 0.001$ |
| **Alanine aminotransferase $> 40$ U/l – No., %** | 20(7.5) | 10(4.6) | 10(20.0) | 0.001 |
| **Albumin $< 40$ g/l – No., %** | 63(23.6) | 19(8.8) | 44(88.0) | $< 0.001$ |
| **Total bilirubin $\geq 26$ μmol/l– No., %** | 6(2.2) | 3(1.4) | 3(6.0) | 0.082 |
| **Direct Bilirubin $\geq 8.6$ μmol/l– No., %** | 7(2.6) | 4(1.8) | 3(6.0) | 0.243 |
| **Indirect Bilirubin $\geq 16$ μmol/l – No., %** | 3(1.1) | 2(0.9) | 1(2.0) | 0.465 |
| **Creatinine $\geq 106$ μmol/l– No., %** | 4(1.5) | 1(0.5) | 3(6.0) | 0.022 |
| **D-dimer $\geq 0.5$ mg/l – No., %** | 19(7.1) | 6(2.8) | 13(26.0) | $< 0.001$ |
| **Radiologic findings – No./total No. (%)** |  |  |  |  |
| bilateral ground-glass opacity or patchy shadowing | 100/231(43.3) | 55/181(30.4) | 45/50(90.0) | $< 0.001$ |
| unilateral ground-glass opacity or patchy shadowing | 95/231(41.1) | 90/181(49.7) | 5/50(10.0) | $< 0.001$ |
| consolidation or fibrous stripes-like shadows | 13/231(5.6) | 4/181 (2.2) | 9/50 (18.0) | $< 0.001$ |
| pneumothorax | 1/231(0.4) | 0/181 | 1/50 (2.0) | 0.216 |
| hydrothorax | 1/231(0.4) | 0/181 | 1/50 (2.0) | 0.216 |
| no radiological abnormality | 36/231(15.6) | 36/181(19.9) | 0 | 0.001 |

Among the 96 patients underwent for lymphocyte subsets detection, decreases in CD4 positive T cell counts were observed in 77.1 % of cases, especially in the severe patients (45, 100%). More patients (53.1%) had decreased CD positive 3 T cell counts, the level of CD8 positive T cells was lower than the normal range in part of patients (34.4%). The average level of haemoglobin was in normal range (135.0 [95.0-165.0]). Few patients had myocardial zymogram abnormality, which showed elevated level of lactate dehydrogenase (LDH), creatine kinase (CK) and creatine kinase isoenzyme (CKMB) in 21.3%, 18.7% and 12.4% of patients, respectively, which indicates a degree of cardiac injury. Few patients presented mild live function abnormality, with elevated level of alanine aminotransferase (ALT) (20 [7.5%]), aspartate aminotransferase (AST) (19 [7.1%]), total bilirubin (6 [2.2%]), direct bilirubin (7 [2.6%]) or indirect bilirubin (3 [1.1%]). Other dysfunctions in kidney and coagulation were rare, with mild increases of creatinine and D-dimer in 1.5% and 7.1% of patients.

Regarding the infection index, 25.3%, 15.9% and 41.5% of patients had higher level of C-reactive protein (CRP), procalcitonin (PCT) and erythrocyte sedimentation rate (ESR), respectively. Furthermore, there are 67 patients detected for cytokines in peripheral blood. Most patients had normal level of IL-2, IL-4, TNF-α and INF-γ, while high level of IL-6 and IL-17A was common in COVID-19 patients (47 [70.1%], 35 [52.2%]).

Of all patients who underwent radiologic examination (X-ray or CT), most of patients presented as pneumonia with bilateral ground-glass opacity (43.3%) or unilateral ground-glass opacity or patchy shadowing (41.1%), co-existed with consolidation or fibrous stripes-like shadows (5.6%). Pneumothorax or hydrothorax occurred rarely (1 [0.4%], 1 [0.4%]). 36 cases (15.6%) had no obvious radiological abnormality.

Additionally, Figure1-3 demonstrates the representative radiologic images of patients with non-severe and a patient with severe COVID-19 in Chongqing.

**Fig.1.**
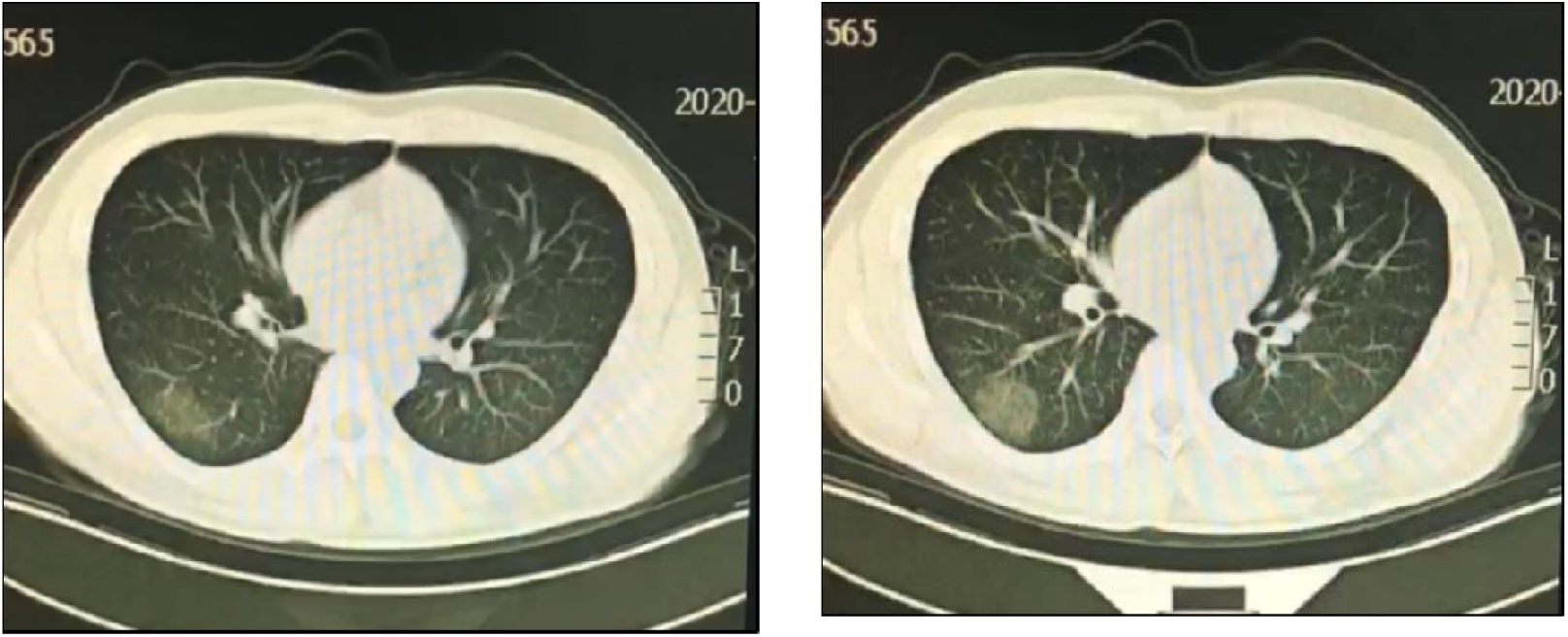
Chest computed tomography of a 56 years old female patient of non-sever COVID-19 in Chongqing on admission.

**Fig.2.**
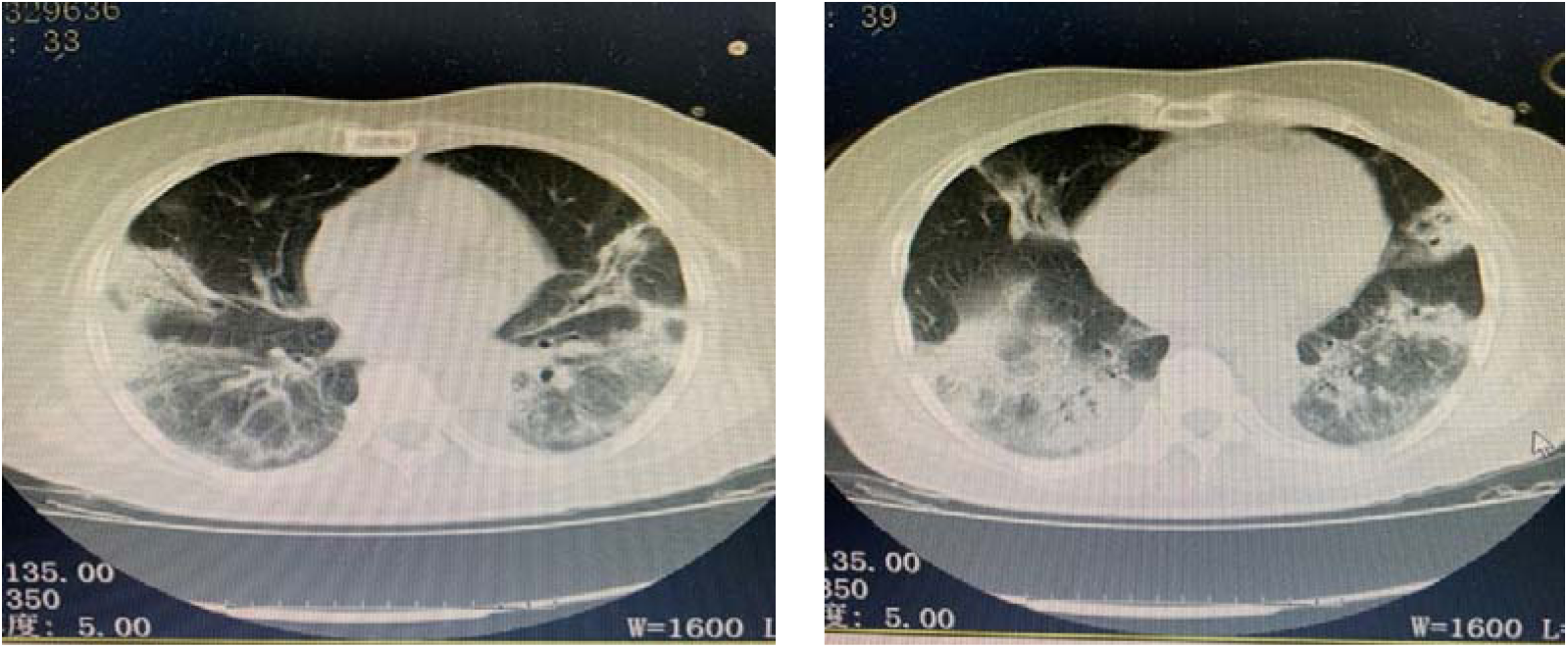
Chest computed tomography of a 65 years old female patient of severe COVID-19 in Chongqing on admission.

**Fig.3.**
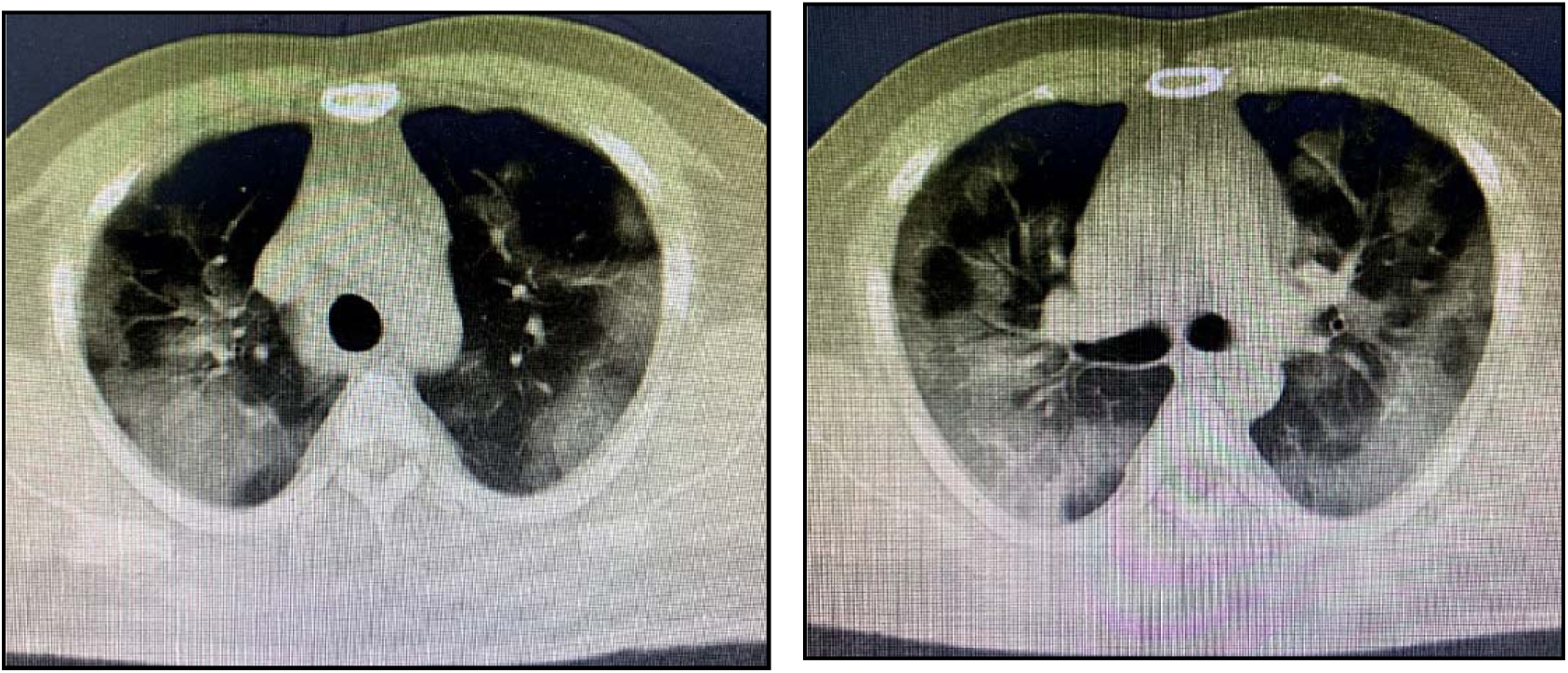
Chest computed tomography of a 39 years old male patient of severe COVID-19 in Chongqing on admission.

### Comparison of non-severe and severe patients

On admission, 217 (81.27%) and 50 (18.73%) patients were categorized into non-severe and severe subgroups, respectively. Compared with non-severe patients, severe patients were significantly older (median age, 71.5 years [IQR, 65.8-77.0] vs 43.0 years [IQR, 32.5-57.0]) and more likely to be male (110[50.7%] vs 39[78.0%]). Compared with non-severe cases, more severe cases are smokers (31[62.0%] vs 22[10.1%]). Severe patients were more likely to have coexisting disorders (15[30.0%] vs 26[12.0%]), including overweight or obesity (14[28.0%] vs 16[7.4%]), hypertension (13[26.0%] vs 7[3.2%]), diabetes (12[24.0%] vs 14[6.5%]), respiratory disease (10[20.0%] vs 15[6.9%]). Respiratory rate, heart rate, and mean arterial pressure did not differ markedly between non-severe and severe cases on admission. No significant differences in the exposure history between the two groups were observed (all *p*>0.05).

Fever and cough were still the most common symptoms in both non-severe (186[85.7%], 155[71.4%]) and severe cases (39[78.0%], 34 [68.0%]). Compared with the non-severe patients, severe patients were more likely to present dyspnea (17[34.0%] vs 26[12.0%]) and confusion (10[20.0%] vs 15 [6.9%]) symptoms. Few severe patients presented with mild symptoms such as myalgia or arthralgia (37[74.0] vs 99[45.6%]), which should be more noteworthy to avoid delayed or missed diagnosis. The rales and wheezes were more common in severe cases as compared with non-severe cases (21 [42.0%] vs 11[5.1%], 10[20.0%] vs 10[4.6%])).

Compared with non-severe patients, most severe patients demonstrated more laboratory abnormalities, including lower level of lymphocytes (46[92.0%] vs 185 [85.3%]), leukocytes (31[62.0%] vs 87[40.1%]), thrombocytes (19[38.0%] vs 42 [19.4%]) and albumin (44[88.0%] vs 19[8.8%]), as well as higher level of D-dimer (13[26.0%] vs 6[2.8%]), C-reactive protein (29 [58.0%] vs 16 [12.5%]), procalcitonin(14[28.0%] vs 10 [9.9%]), lactate dehydrogenase (18[36.0%] vs 39[18.0%]), creatine kinase (18[36.0%] vs 32[14.7%]), myoglobin (7[33.3%] vs1 [1.8%]) and troponin (3[14.3%] vs 0[0%]). As for lymphocyte subsets, significantly more severe patients had lower level of CD4 positive T cells and CD3 positive T cells (45[100.0%] vs 29[56.9%], 31[68.9%] vs 20[39.2%]). More severe cases had lower level of CD8 positive T cells (20[44.4.0%] vs 13[25.5%], *p*=0.051), while no significant difference was shown. Further comparison the inflammatory cytokines between severe and non-severe patients showed that the level of IL-6, IL-17A and TNF-α was remarkably elevated in severe patients (32[84.2%] vs 15[51.7%], 25[65.8%] vs 10[34.5%], 17[44.7%] vs 5[17.2%]).

### Treatment and complications

All patients were treated in isolated wards, among whom critically severe patients were treated in intensive care unit with negative pressure wards. The treatment, complications and outcomes are shown in Table 3. All patients received antiviral therapy (267, 100%), including interferon, opinavir, arbidol, ribavirin. Most non-severe patients received antiviral therapy only for pharmacotherapy. Few non-severe patients (7, 3.2%) with bacterial infection evidence received empirical antibiotic treatments (moxifloxacin and others), while more severe patients received antibiotic treatments (36, 72.0%). 5 severe patients received antifungal therapy (caspofungin) (5, 10.0%). None of non-severe patients received systemic glucocorticoid therapy, while a portion of severe patients (38, 76.0%), without obvious immune deficiency, received systemic corticosteroid therapy, usually methylprednisolone 40-80mg for 3-5 days. There is only one critically severe patient who received 120mg methylprednisolone. A total of 35 patients (70.0%) received immunopotentiators (thymalfasin 1.6mg/day) or immunoglobulin treatments and 13 severe patients (26.0%) received vasopressors for shock rescue.

**Table 3.**
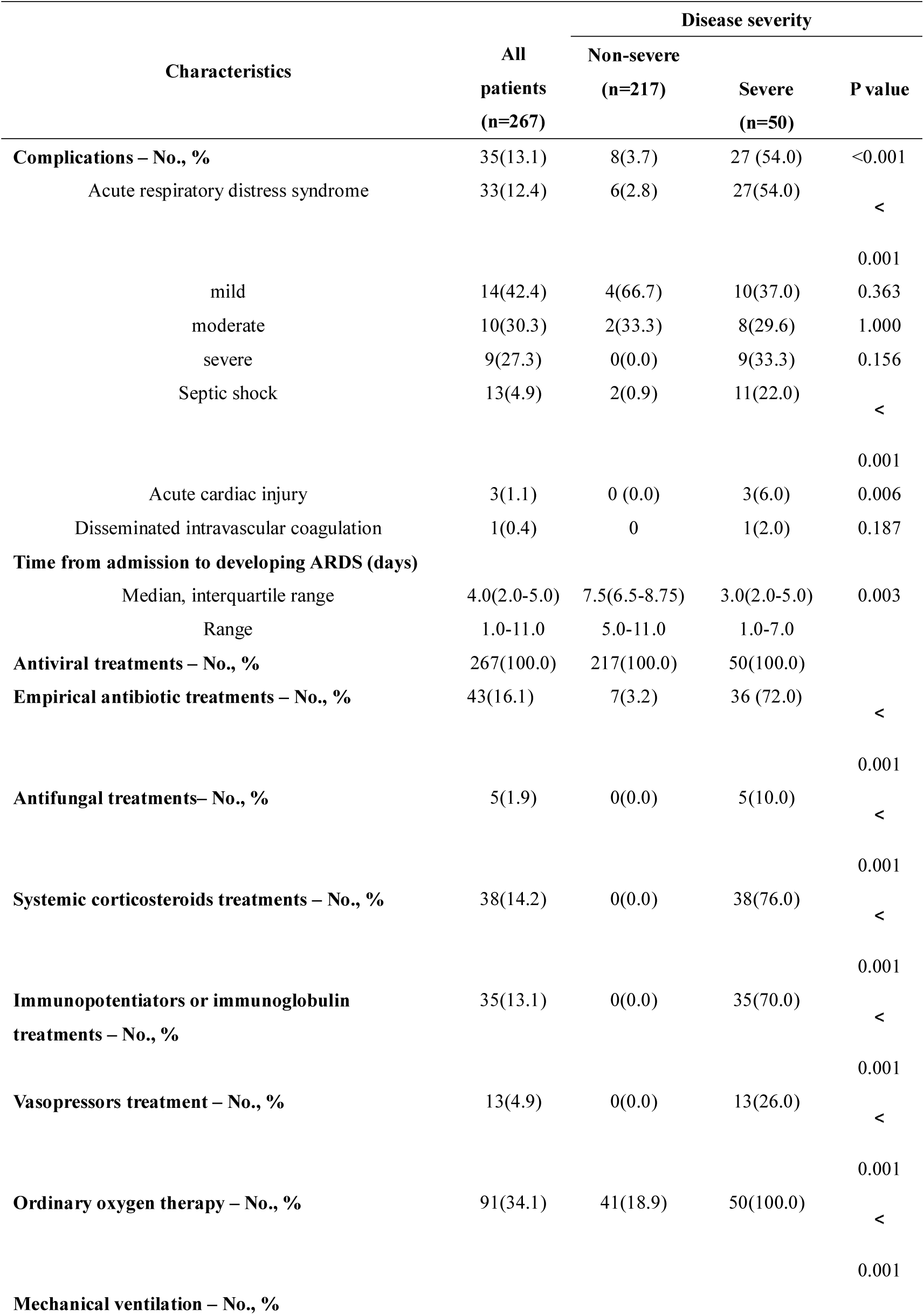
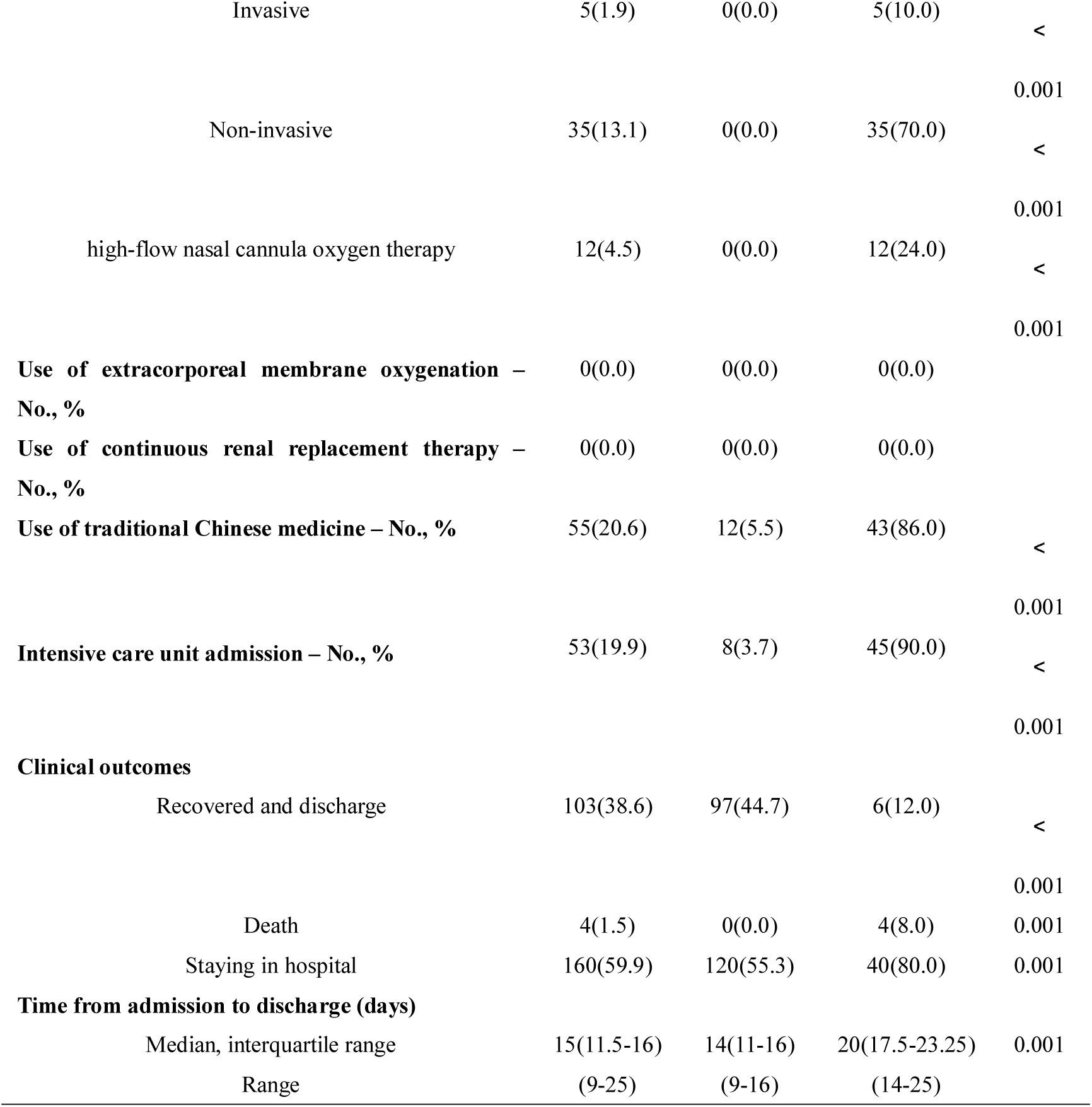
Complications, treatment and outcomes of COVID-19 patients in Chongqing.

Few non-severe patients received ordinary oxygen therapy, delivered by nasal cannula or face mask at 1-4 liters per minute (41, 18.9%). Oxygen therapy including invasive mechanical ventilation, non-invasive mechanical ventilation, high-flow nasal cannula oxygen therapy (HFNC) was adopted only in severe patients with respiratory failure (5[10.0%], 35[70.0%], 12[24.0%]). It is worth noting that 3 critically severe patients received invasive mechanical ventilation in prone position, and the oxygenation improved remarkably. Moreover, traditional Chinese medicine was adopted to most of severe patients (43,86.0%). Currently, none of patients received extracorporeal membrane oxygenation (ECMO) therapy, kidney replacement therapy or convalescent plasma therapy. However, 3 critically severe patients are going to receive ECMO after professional assessment. And, two discharged COVID-19 patients promised to donate convalescent plasma for critically severe patients’ further treatment. As compared with non-severe cases, severe cases suffered higher rates of any complications (27[54.0%] vs 8[3.7%]) during hospital admission. The most common complications were ARDS (33, 12.4%), secondary to shock (13, 4.9%) and acute cardiac injury (3, 1.1%), especially more common for severe patients (ARDS, 54.0%; septic shock, 22.0%; acute cardiac injury, 6.0%). Notably, in our study, all of the critically severe cases are complicated with ARDS of different severity, and 4 death cases died of severe ARDS.

### Clinical outcomes

By February 16, 2020, the health commission of Chongqing announced that among 551 confirmed COVID-19 patients in Chongqing, 207(37.57%) patients were discharged, 5 patients (0.91%) were died and 339(61.52%) patients were still in hospital for treatment, lower than that of Hubei province and mainland China including Hubei province (Fig.4). In our study, 103(38.6%) patients had been discharged and 4(1.5%) patients had died, and all other patients were in hospital. The percentages of patients being admitted to the ICU was 19.9%. The median length of hospital of COVID-19 patients is 15 days (IQR, 11.5-16.0), with longer length of hospital of severe patients than that of non-severe patients (20 days [IQR, 17.5-23.25], 14 days [IQR, 11.0-16.0]).

**Fig.4.**
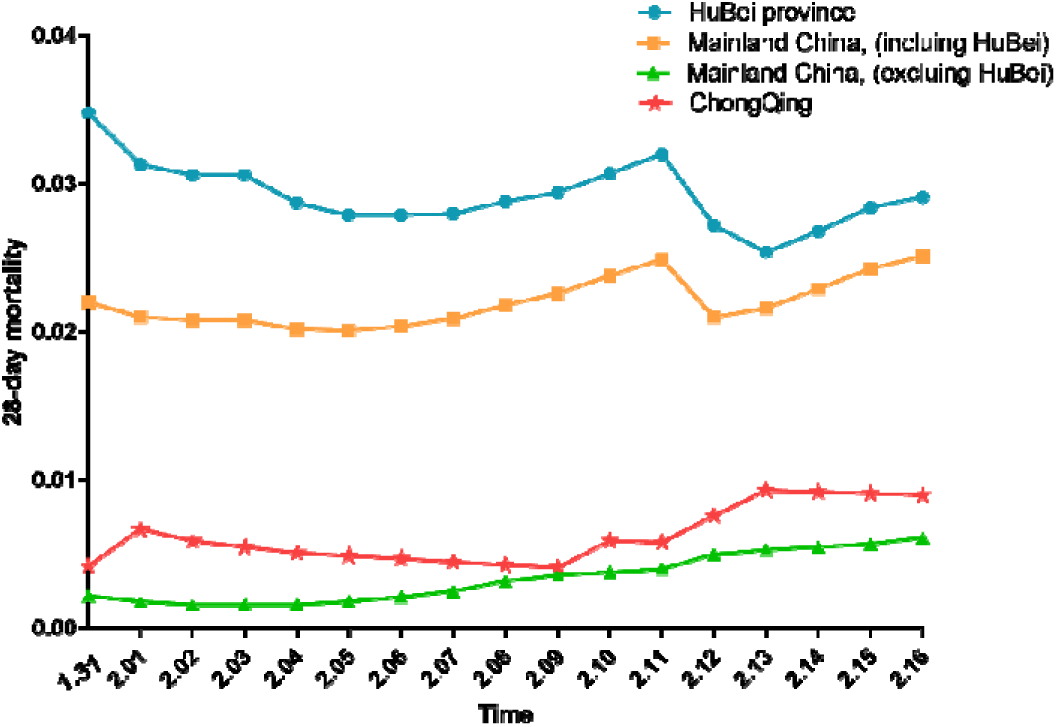
Comparison of 28-day mortality among Hubei province, mainland China including Hubei province, mainland China excluding Hubei province and Chongqing municipality.

For severe patients, the duration from admission to developing ARDS is only 3 days (IQR, 2.0-5.0), and the 28-mortality of severe patients was 8.0% (4/50), with much higher when complications occurred (14.8%, [4/27]). Notably, the 28-mortality of critically severe patients complicated with severe ARDS is considerably as high as 44.4% (4/9). Therefore, early diagnosis and intensive care of critically severe COVID-19 cases will be considerably essential to reduce complications and mortality.

## Discussion

In December 2019, COVID-19 firstly occurred in Wuhan, Hubei Province, China. The disease has rapidly spread from Wuhan to worldwide^1^. On January 30, the World Health Organization has declared the COVID-19 outbreak as a public health emergency due to this rapid infection spreads. However, as one of the major cities imported passengers from Wuhan in China^9^, the regional characteristics of COVID-19 in Chongqing municipality were not reported so far. Our presents study, to our knowledge, firstly portrayed the epidemiological, demographic, clinical, radiological, laboratory characteristics, treatment regimens and clinical outcomes of COVID-19 patients in Chongqing municipality, China. And we further compared the numerous differences between severe and non-severe patients in Chongqing municipality. The 28-day mortality of COVID-19 patients from 3 designed hospitals of Chongqing municipality is 1.5%, approximately consistent with the official fatality rate 0.91%, lower than the 28-day mortality of mainland China including Hubei province (2.51% [1770/70548]), reported by national official statistics as for February 16th, 2020 Recently, there are some studies described the cases of hospitalized patients with COVID-19 in Wuhan or worldwide. First study including 41 cases of COVID-19 in Wuhan, in which most patients had a direct history of exposure to Huanan Seafood Wholesale Market of Wuhan, with high mortality rate (6/41, 15%) in this study^5^. An extended descriptive study from 99 cases in the same hospital further revealed the epidemiology and clinical characteristics of the COVID-19 in Wuhan^6^. Another study from 138 hospitalized COVID-19 patients in Wuhan found that 26% of patients received intensive care unit (ICU) care and further compared the differences between severe cases received ICU care with non-severe cases who did not receive ICU care^7^. Two more researches including COVID-19 cases based on the big data from the whole country delineated the clinical and epidemiological characteristics of COVID-19 in nationwide^8,13^. The clinical courses and outcomes of critically ill COVID-19 patients was recently reported^14^.

In our study, the majority critically severe patients are older and male, consistent with gender difference in previous researches^6,13,14^. The high susceptibility of males is similar to MERS-CoV and SARS-CoV infection^15,16^. Although, only few of patients in Chongqing are smokers, more severe cases are smokers. Previous study demonstrated that the gene expression of ACE2, receptor of 2019-nCov, is significantly higher in smoker’s lung. ACE2 gene is expressed in specific cell types related to smoking history and actively expressed in remodeled AT2 cells of smokers. Thus, smoking history may indicate a poor prognosis due to its different infection paths with non-smokers^17^.

Compared with patients who had a definite direct exposure to Huanan seafood market in Wuhan, the epidemics in Chongqing has transmitted to the coexistence of imported cases and local sporadic or clustered cases. As the strong epidemic control in Hubei was implemented, clustered cases or family aggregation cases gradually compose the majority of confirmed cases in Chongqing. Although small part of patients in Chongqing had coexisting chronic disease, relatively less frequent than that of patients in Wuhan^6^, severe patients were more likely to suffer from underlying comorbidities (diabetes, obesity, cardiovascular and cerebrovascular diseases), resulting in the deficiency in innate and adaptive immunity of these patients. Among the common comorbidities, we found a significant portion of patients in Chongqing suffered from overweight and obesity, which is more common in severe patients. Obesity-induced chronic inflammation status is well documented to contribute to the progress of multiple diseases^18^. Large epidemiological studies have shown that BMI was associated with increased risk of acute respiratory distress syndrome (ARDS) and length of hospital stay, which has already been used in lung injury prediction score (LIPS) to help physicians identify at-risk patients for developing ALI^19^. Therefore, elderly male patients with a history of smoke and underlying comorbidities are at increased risk of becoming critically severe if they are suffered from COVID-19 infection, which deserves more attention and intensive care treatment.

As the sequence of 2019-nCoV showing 79.0% nucleotide identity with the sequence of SARS-CoV, and 51.8% identity with the sequence of MERS-CoV^20^, the clinical characteristics of COVID-19 infection bear resemblance to SARS-CoV and MERS-CoV infection. Dominant symptoms at onset of COVID-19 in Chongqing were still fever, dry cough, fatigue, dyspnea, and myalgia or arthralgia is more common in the Chongqing patients. The absence of fever, especially hyperpyrexia, may increase the difficulty to identify and diagnosis COVID-19 in clinical practice if too much attention was given to fever detection only. Moreover, in our study, a significant proportion of patients in Chongqing initially developed with atypical symptoms, such as upper respiratory tract symptoms (eg, nasal congestion and pharyngalgia) and gastrointestinal symptoms (eg, anorexia, diarrhea and nausea), and even had no obvious symptoms, which is inconsistent with previous study reporting upper respiratory tract and intestinal signs and symptoms are rare in Wuhan province^5^. Therefore, the atypical patients and asymptomatic carriers deserves more attention to avoid delayed or missed diagnosis. Moreover, rales and wheezes are scarcely noted when lung auscultation was performed, due to the lack of sputum or mucus, especially for non-severe patients. Further comparison between the severe and non-severe patients found that severe patients are more likely to suffer from dyspnea and confusion symptoms on disease onset, which may suggest severe complications such as respiratory failure or septic shock, and is of great value of early identification of critically severe cases. The median durations from symptoms onset to laboratory diagnosis and hospital admission were 4 days and 7 days, with obvious longer time for severe cases, which may lead to the poor outcome due to delayed medical interventions. In general, COVID-19 in severe cases progresses rapidly to lethal complications (ARDS, septic shock, acute cardiac injury, refractory metabolic acidosis and so on), even leading to death. Our study demonstrates that the median interval from symptoms onset to ARDS of severe cases was much shorter than that of non-severe cases. It has been demonstrated that poor populations always endured a disproportionate burden of disease and death from infectious diseases like influenzas^21^. A great number of death cases in our study were poorly-educated elders with chronic comorbidities, their illness deteriorated rapidly to severe ARDS within several days, presented as suddenly occurred severe hypoxaemia, malignant arrhythmia, cardiac and respiratory arrest, unfortunately death, despite of timely rescue including invasive ventilation and other advanced medical interventions. Therefore, early diagnosis and management of critically severe COVID-19 cases, especially poorly-educated elderly male with chronic comorbidities, should be vigorously advocated.

Regarding to laboratory tests, consistent with most reports, peripheral lymphocytopenia is the most crucial and common feature for COVID-19 infection. In our study, most patients suffered from varying degrees of lymphocytopenia, especially for critically severe patients. Lymphocytopenia is also a prominent feature for patients with SARS-CoV, H1N1, MERS-CoV virus infection^22-25^. Progressive lymphopenia is reported to occurred early in the course of SARS and reached its lowest point in the second week in most patients, then, lymphocytes count commonly recovered in the third week^25^. In terms of lymphocyte subsets, the counts of total T cells, CD4 positive and CD8 positive T cells had reduced in COVID-19 patients on admission. Counts of total T cells and CD3 positive T cells were significantly lower among severe patients, accompanied by an obvious declining tendency in CD8 positive T cells, indicating an adverse outcome. Pathological findings from the biopsy of COVID-19 death patient implied that overactivation of T cells, manifested by increase of highly proinflammatory Th17 and cytotoxic CD8 T cells, partially accounts for the severe immune injury in death patient^26^. The phenomenon that T cell counts reduced and functionally exhausted has been reported in previous study, which indicates high risk for further deterioration of COVID-19^27^. In our study, a progressive decline of lymphocytes and CD4 positive T cells was observed during the deterioration of illness. Thus, the dynamic profile of lymphocytes and lymphocyte subsets are of great value for disease progression and outcomes prediction. Early studies have shown that 2019-nCoV infection increased the release of both inflammatory and anti-inflammatory cytokines from T-helper-1 (Th1) and T-helper-2(Th2) cells, differing from SARS or MERS-infection induced proinflammatory cytokines secretion^28,29^. Our study demonstrated that pro-inflammatory cytokines IL-6, IL-17A were elevated in mostly patients, with significant higher level of IL-6, IL-17A and TNF-α in severe patients, indicating an underlying relationship between pulmonary inflammation and lung damage in 2019-nCoV patients. So far, the cytokines storm’s effects on viral pneumonia are considerable complex and their clinical roles in severe lung injury have not been extensively documented, therefore, further investigations are needed to elucidate immune and inflammation response in 2019-nCoV pathogenesis, which is of crucial importance for efficient treatments. Few of patients presented as elevated levels of infectious parameters, but other abnormalities were less common. While, severe patients obviously manifested more prominent abnormalities, suggesting multiple organ dysfunction and poor outcomes. Additionally, severe cases suffered from lower albumin and hemoglobin, suggesting the importance of nutrition supportive treatments. As for radiologic presentations, more attention should be paid to the cases without obvious radiological abnormalities on admission, so as to avoid the miss or delayed diagnosis. Therefore, it is crucial important to take symptoms, laboratory findings, radiologic findings into account to make an integrated and thorough judgment. The MuLBSTA score, an efficient early warning model for predicting mortality in viral pneumonia^30^, needs further validation in the future practice.

Currently, no specific treatment available has been advocated for coronavirus infection. As most of patients in Chongqing were non-severe, antiviral therapy was commonly adopted among these patients. Empirical antibiotic therapy was merely administered to few patients with bacterial infection evidence, and ordinary oxygen therapy was supplied only if hyoxemia occurred. As for the severe cases, comprehensive treatment is important. Mechanical ventilation is the main respiratory supportive treatment for critically ill patients, which should be administered as soon as possible if the normal oxygenation cannot maintain. Treatments were focused on supportive therapy, which focuses on limiting further lung damages through a combination of lung-protective ventilation to prevent ventilator-associated lung injury and conservative fluid therapy to prevent the hyperpermeability of alveolar endothelial and epithelial barriers and promote lung edema resorption.

Although, systematic corticosteroid treatment is not routinely recommended for COVID-19 patients^31^, while based on the pathological findings of COVID-19 biopsy that pulmonary edema and hyaline membrane formation^26^, low dosage of intravenous methylprednisolone (usually 20-80mg for 3-5 days) was still provided to some severe patients at early stage of the illness to suppress lung inflammation and hasten radiographic improvement. Immunopotentiator and γ-immunoglobulin should be administered to enhance immune responses and pathogen clearance based on the patient’s condition. Notably, traditional Chinese medicine formulae was extensively utilized in critically severe patients after precise and professional evaluation. Two discharged COVID-19 patients promised to donate convalescent plasma for critically severe patients’ further treatment. Previous Meta-Analysis reported that patients with H5N1 influenza pneumonia who received influenza-convalescent human blood products may have experienced a reduction in the risk for death^32^. Convalescent plasma may reduce the mortality of SARS coronavirus and severe influenza infection^33^. On the contrary, high-titre anti-influenza plasma was reported to confer no significant benefit over non-immune plasma for treating patients with severe influenza A^34^. Transfusion of convalescent plasma into 84 patients with Ebola virus disease was not associated with a significant improvement in survival^35^. Therefore, it is still too early to judge the efficiency of these therapy, much more studies are still needed In summary, our study firstly demonstrated the regional disparity of COVID-19 in Chongqing and further thoroughly compared the differences between severe and non-severe patients in Chongqing municipality, China. The 28-day mortality of COVID-19 patients from 3 designed hospitals of Chongqing municipality is 1.5%, approximately consistent with the official fatality rate 0.91%, lower than that of Hubei province and mainland China including Hubei province. Early isolation, early diagnosis, early management, combined with the strong epidemic control in Hubei and the effective prevention of local cluster outside Hubei might collectively contribute to the marked reduced 28 day-mortality in Chongqing municipality.

However, for severe patients, the 28-mortality was relatively high, with much higher when complications occurred. Notably, the 28-mortality of critically severe patients complicated with severe ARDS is considerably as high as 44.4%. Thus, early diagnosis and intensive care of critically severe COVID-19 cases, especially those combined with ARDS, will be considerably essential to reduce mortality.

## Data Availability

All data referred to in the manuscript are availability.

https://pan.baidu.com/s/12c1bSopgxC9EbIovynpCvw

## Acknowledgements

Daoxin Wang, Jin Tong, Jianglin Xiang designed the research; Di Qi, Xiaofeng Yan, Xumao Tang, Junnan Peng, Longhua Feng, Guodan Yuan, An Zhang, Yaokai Chen, Jing Yuan, Xia Huang, Xianxiang Zhang, Peng Hu, Yuyan Song, Chunfang Qian, Qiangzhong Sun collected the data; Di Qi, Xumao Tang, Junnan Peng,Qian Yu performed data analysis; Di Qi, Xiaofeng Yan performed manuscript writing; Di Qi,Xumao Tang, Junnan Peng, Qian Yu contributed to study advice. Daoxin Wang, Jin Tong, Jianglin Xiang directed the study and participated in the review. Di Qi, Xiaofeng Yan, Xumao Tang contributed equally.

## Ethics

This research was approved by the institutional ethics board of the Second Affiliated Hospital of Chongqing Medical University (No.2020-09), Chongqing public health medical center (No.2020-015-01-KY), Chongqing three gorges central hospital (No.2020-13) and Qianjiang central hospital of Chongqing (No.2020-07).

